# Strengthening Laboratory Management Towards Accreditation in Tanzania: the 13 Years of Remarkable Revolutions in Laboratory Quality Management System

**DOI:** 10.1101/2025.02.27.25322997

**Authors:** Peter Richard Torokaa, Charles Massambu, Jacob Lusekelo, Reginald Julius, Alex S. Magesa, Mtebe Majigo, Agricola Joachim, Maria E. Kelly, Alice Kyalo, Nyambura Moremi

## Abstract

**Introduction:** The increase in demand of laboratory services and recent advancements in laboratory technologies underscore the importance of quality laboratory services in delivering accurate, reliable, and timely test results. As a strategy to ensure quality laboratory services in the country, Tanzania started to implement Strengthening Laboratory Management Towards Accreditation (SLMTA) program in 2010. We describe the revolutions made in laboratory quality management system across the 13 years of SLMTA implementation highlighting the impact and achievements attained following the modification of the generic SLMTA approach.

**Methodology:** We reviewed, described and summarized the 13 years of SLMTA implementation in Tanzania and the quality performance of medical laboratories enrolled into the program. The quality performance was evaluated based on the increase in number of SLMTA laboratories accredited under ISO 15189 by comparing the laboratories enrolled during the generic versus modified SLMTA program.

**Results:** Out of 138 SLMTA laboratories assessed by 2023 using the SLIPTA checklist, 81 (58.7%) scored 3 stars and above were enrolled in the accreditation process. Of the 81 enrolled laboratories, 50 (61.7%) achieved ISO 15189 accreditation status by 2023. There was a significant increase in number of accredited laboratories during the five years (2018-2023) of modified SLMTA compared to the eight years (2010-2018) of generic SLMTA (47/64 vs. 3/17, *p*= 0.021). In the context of level of healthcare delivery, most of the accredited laboratories (n=47/50, 94%), belonged to the district and above.

**Conclusion:** To the best of our knowledge, this is the first report of the modified SLMTA approach with remarkable results. The modified SLMTA program significantly improved the status and number of accredited laboratories in Tanzania. While the attained milestone demonstrates the SLMTA program’s effectiveness in strengthening quality of laboratory services, the significant achievements underscore the essence of monitoring, evaluation and learning of the implemented programs, and most importantly the need for strategies that contextualize and tailor the program for sustainability.

## Introduction

Quality laboratory services play a crucial role in the delivery of health services in the country by ensuring accurate, reliable and timely test results. Quality laboratory results are key in confirming the clinical diagnosis for guiding patient management, disease prevention and outbreak confirmation (1). Moreover, laboratory results have been valuable in informing health policy and providing guidance in the development of treatment guidelines, and monitoring of disease trends as applied in surveillance systems. By establishing a laboratory network of ISO 15189 accredited laboratories that have well established, documented and implemented Quality Management System (QMS), we ensure timely, accurate and reliable laboratory results for decision making. Therefore, strengthening of laboratory services through quality improvement programs and accreditation remains critical in order to safeguard individual and community health (2,3).

The role of laboratory services in clinical and public health settings has been profoundly influenced by recent developments in biomedical sciences notably in the detection and management of infectious diseases (1). In the context of pandemic preparedness, detection, response and recovery, the International Health Regulations (IHR) clearly emphasizes the importance of strengthening laboratory systems and networks in support of national and regional disease surveillance for early detection and response to health emergencies including disease outbreaks (4).

In support of initiative to enhance the quality of laboratory services to improve patient care (5), the President’s Emergency Plan for AIDS Relief (PEPFAR) initiative through the U.S. Centers for Disease Control and Prevention (CDC) in collaboration with the World Health Organization (WHO) regional offices for Africa (WHO AFRO) developed and launched the Strengthening Laboratory Management Towards Accreditation (SLMTA) in 2009 (6). The goal of SLMTA program was to improve the quality and efficiency of laboratory services in resource-limited settings, enabling laboratories to achieve and sustain international accreditation standards in limited settings including Tanzania. The SLMTA program provides the how-to with training and mentorship (6). Two years following the implementation of SLMTA i.e., in 2011, the WHO-AFRO in collaboration with African Society for Laboratory Medicine (ASLM) realized the need to measure and acknowledge the level of implementation and improvement of SLMTA. This idea led to the birth of a sister program namely Stepwise Laboratory Improvement Process Towards Accreditation (SLIPTA) (7). As a complete package, the SLMTA/SLIPTA program came as a competency-based solution and stepwise approach towards the implementation of the quality management system in clinical, reference and public health laboratories within African member states (8).

Tanzania adopted the SLMTA and SLIPTA programs in 2010 and 2013, respectively. The 13 years of SLMTA implementation in Tanzania has positively impacted the laboratory space and positioned Tanzania among the top three countries with highest percentage of accredited during the 6^th^ Biennial SLMTA Symposium in 2023. In this article, we describe the 13 years of implementing the SLMTA program in Tanzania, chronicling the progress, modification and milestones achieved throughout the process. Tanzania began with the introduction of the SLMTA program, which aimed at enhancing QMS in laboratories across the country. Over time, the program evolved and adapted to better suit the local context and needs, leading to significant improvements in laboratory accreditation rates. This transformation and revolution marked a pivotal point in Tanzania’s healthcare landscape, showcasing the resilience and dedication of those involved in the process. The positive impact of these efforts are evident in the increased number of accredited laboratories and the overall enhancement of healthcare services, contributing to better health outcomes for the community.

## Methodology

### SLMTA implementation (2010-September 2018) using the generic approach

We reviewed the implementation of the adopted generic SLMTA program in Tanzania from 2010 to September 2018. This program lasted between 12 and 18 months, with three cycles of training, mentorship and on-site supervision, and monitoring which included working on improvement projects (Fig 1).

**Fig 1.**
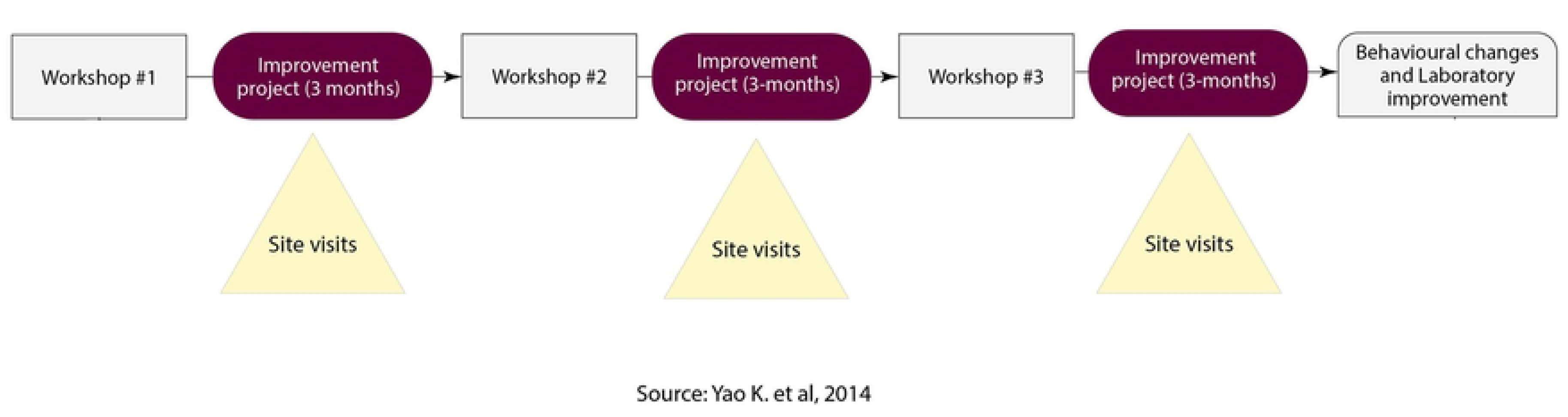
The generic SLMTA implementation approach.

In review of the generic SLMTA approach, it was designed to enable participants develop ten critical competencies for laboratory managers in ten key areas such as productivity, work area, inventory, procurement, equipment maintenance, quality assurance, specimens, laboratory testing, test result reporting, and document and record control. Furthermore, the competency areas comprised of 66 laboratory management activities and 45 training activities. Under the generic model, the training part was delivered through workshops, whereby three workshops were conducted throughout the program duration each lasting for five days (45 hours), incorporating 45 instructional activities and 66 laboratory management activities (9).

The three five-day workshops were held at a three-month interval with a goal of imparting the knowledge and skillset to laboratory managers and quality officers as per curriculum package. Workshop participants were given tools such as laboratory management framework, detailing organizational structure, roles, and responsibilities. Assessment checklist which provides a structured approach for internal audits, covering all aspects of laboratory operations, and SLMTA toolkit that includes modules on leadership, process control, documentation, and safety, among others, providing laboratories with the necessary resources to enhance their quality management practices and comply with ISO 15189 standard. These tools collectively supported laboratories in achieving and maintaining high-quality standards, ensuring reliable and accurate diagnostic results.

For the assessment checklist, initially (2010-2012) the baseline audits were conducted and evaluation of the SLMTA program was assessed using the WHO-AFRO accreditation checklist, until the year 2013 where the SLIPTA checklist was adopted.

In addition, participants were also provided with SLMTA toolkit that outline best practices and improvement projects that either incorporate lessons learnt from previous workshops or address common nonconformities identified during baseline audits (10).

To ensure the effective implementation of the SLMTA program, the Tanzanian SLMTA Supervisory Team conducted supportive supervision visits. These visits aimed to oversee the execution of improvement projects, provide clarity on complex issues, and offer guidance. This included translating workshop content taught in English into Kiswahili, the national language, to ensure better understanding and implementation of the SLMTA program (10).

In terms of assessment modalities, each enrolled laboratory underwent audits at the start (baseline assessment) and at the end of the program (exit assessment). These audits used a comprehensive checklist to evaluate compliance and performance. By comparing the baseline and exit scores, as well as star ratings (11), the program’s impact on laboratory function and quality was measured (9). This method allowed for a clear assessment of the progress and effectiveness of the SLMTA program in improving laboratory standards.

Furthermore, we described other key aspects of implementation of SLMTA program from the point of enrolment of the laboratory into the program up to attaining international accreditation under ISO 15189. Such aspects include selection and enrolment of the laboratories into SLMTA program, Training of mentors and assessors, Evaluation of the SLMTA program and enrolment of these laboratories into the accreditation program.

### Selection and enrolment of laboratories to SLMTA program

The Ministry of Health (MoH) through the Laboratory Technical Working Group (TWG) identified the minimum criteria for selecting laboratories for the program. The criteria included the number of qualified laboratory staff according to the healthcare delivery level, basic knowledge of quality management system, the facility infrastructure and participation in an external quality assessment program (10). After the initial evaluation, a list of pre-selected laboratories was provided to certified assessors for physical visits and verification of the information provided. The final assessor’s report was then made available for the decision to either enrol or drop the respective laboratories.

### Training of Mentors and Assessors

Under the technical assistance from Clinical Laboratory Standard Institute (CLSI), the MoH provided a five-day training to laboratory mentors. A total of 89 mentors were trained with a goal of capacitating mentors with knowledge and skillset to mentor and provide technical assistance to laboratories implementing the SLMTA program. The implementation of SLMTA program was also backed up by the African Society for Laboratory Medicine (ASLM) where a total of 37 assessors were trained for a five-day program. The training was based on the assessment of laboratories using SLIPTA checklist and the fulfillment of ISO 15189 standard requirements. The participants for mentors and assessor’s training were selected from laboratories that attained three stars and above or from ISO 15189 accredited laboratories.

### Assessment and grading of SLMTA laboratories

Assessment reports (completed SLIPTA checklists and table of non-conformities) were mandatory for assessment and grading. Two different evaluation assessments (internal and external assessment were conducted at the end of the SLMTA program circle. The internal assessment was coordinated and organized by the MoH using the in-country trained and certified SLIPTA auditors. The findings from this assessment were used to select laboratories for external assessment organized and coordinated by ASLM. On the other hand, findings from the MoH and ASLM assessment were used to guide the selection of laboratories enrolment into the accreditation program. Using the SLIPTA checklist, the laboratories were graded on a scale of 0 to 5 stars, indicating the level attained in establishing, documenting and implementing the QMS (11) Table 1.

**Table 1.**
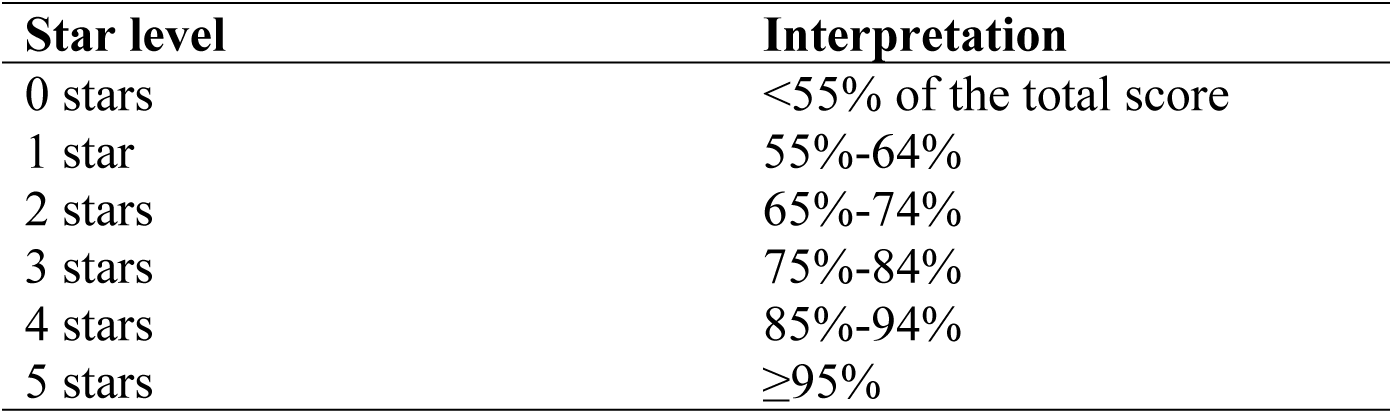
Description of star levels and interpretation from SLIPTA checklist.

| Star level | Interpretation |
| --- | --- |
| 0 stars | <55% of the total score |
| 1 star | 55%-64% |
| 2 stars | 65%-74% |
| 3 stars | 75%-84% |
| 4 stars | 85%-94% |
| 5 stars | ≥95% |

### Enrolment into the accreditation process, the fate of SLMTA program

According to WHO-AFRO SLIPTA guidelines, laboratories that attain 4 or 5 stars may apply for international accreditation. However, laboratories graded below 4 stars can also apply for accreditation given the availability of resources and management commitment (12).

Tanzania, as a member of the Southern African Development Community (SADC), utilizes the SADC Accreditation Service (SADCAS) as its service provider for laboratory accreditation. Under this arrangement, the MoH conducted the in-country mock assessment using in-country assessors to gauge the laboratories prior to the SADCAS assessment. Once the laboratory passes the mock assessment, it was deemed ready, and officially recommended to apply for SADCAS accreditation. Throughout this process, the MoH’s National Laboratory Quality Officer served as a liaison officer between SADCAS and recommended laboratories, facilitating communication and coordination. Moreover, the liaison officer oversaw all the procedures related to the application process, including recommendations, rejections, and periodic surveillance.

### SLMTA implementation (October 2018 to 2023) using in-country modified approach

Towards the end of the year 2018, eight years after the SLMTA adoption, the in-country MoH laboratory TWG evaluated the entire program to identify strengths and gaps with an overall objective of improving the program (13). Among the identified gaps, the critical ones included the challenges in performing root cause analysis, corrective action, internal audit and method verification (13). As a plan to bridge these gaps the in-country MoH laboratory TWG team proposed a modification of the generic SLMTA program to ensure the participants master the areas that were identified as gaps.

### Modified areas

The modification of the approach included the three-week intense training of the four problematic areas (root cause analysis, corrective action, internal audit and method verification) immediately after the baseline assessment (Fig 2).

**Fig 2.**
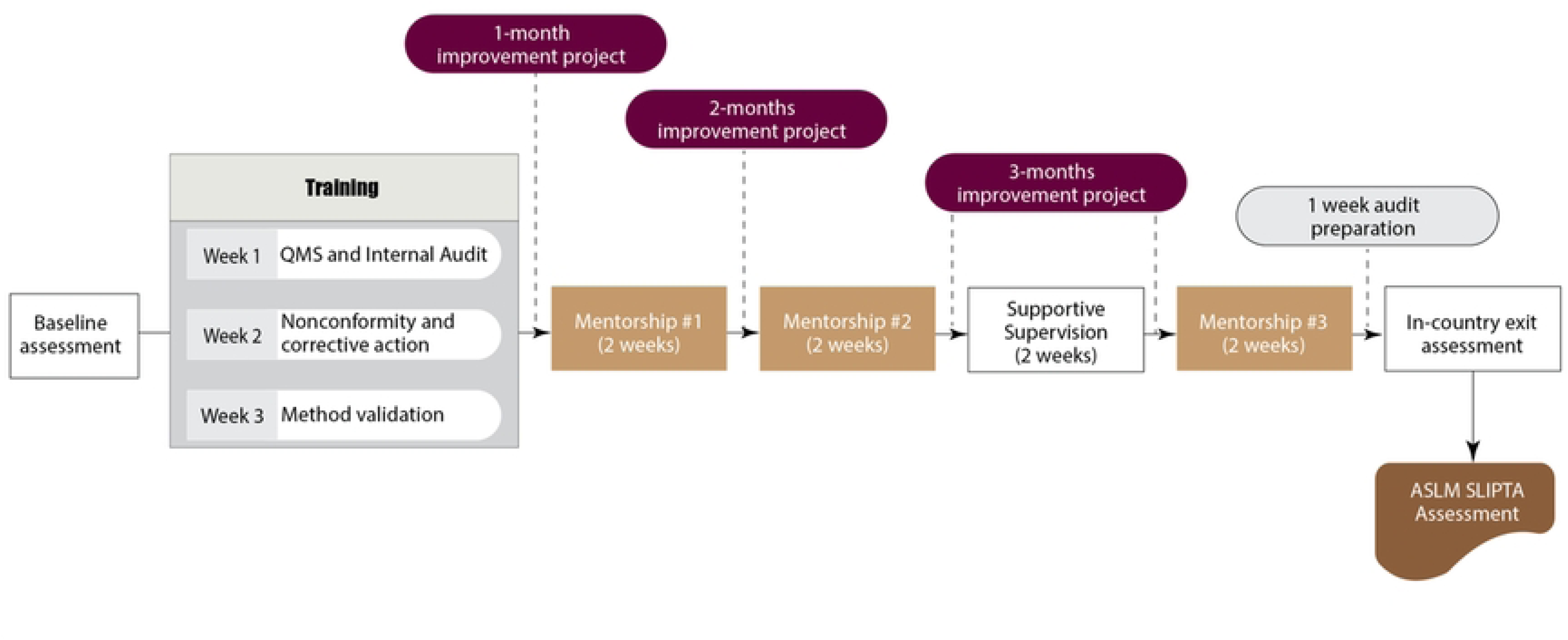
In-country modified SLMTA approach.

Apart from the intense training of the four problematic areas described, the other modification included shortening the SLMTA program duration to a maximum of 9 months from the previous 12 to 18 months (Fig 2). The QMS documents were standardized into templates that laboratories could customize. The developed documents were in line with ISO 15189 requirements and included the Quality Policy Manual, Sample Collection Manual and Safety Manual. Additionally, supportive documents such as forms for documentation and records that provided evidence of QMS implementation, were also standardized.

During site visits, supervision focused on overseeing the practical application of the skills acquired from the intense training on the problematic areas as well as monitoring the execution of improvement projects (like the generic approach). Selection of laboratories was another area that was modified. To address the challenge of minimal engagement and lack of ownership of SLMTA laboratories by the hospital management teams, the MoH opted to advertise the program for the interested health facilities to apply. However, the criteria for the enrolment of laboratories into SLMTA remained the same as in generic approach.

The modified approach was put in place in October 2018, and all laboratories that had been in SLMTA program for the past eight years but still in either zero or one star were enrolled to piloting phase of the modified SLMTA approach. The annual SLMTA meeting was initiated to monitor, evaluate and learn the progress of the modified approach. Members of the meeting included stakeholders from both public and private partnerships. As a follow-up to the discussion of best practices and areas for improvement, the annual SLMTA and accreditation plan for the subsequent year was developed. The annual plan included all QMS matters.

Regarding the accreditation process, a slight modification was also done by adding the in-country self-assessment before the final ASLM SLIPTA assessment. Before the completion of the 9 months of modified SLMTA program, the in-country three certified assessors (one team lead and two technical assessors) conduct a two-day assessment. The laboratories that attained 2 stars and above were subjected to another one-week mentorship before the final ASLM SLIPTA assessment. After the final ASLM SLIPTA assessment, all laboratories that attained 3 stars were enrolled to the accreditation process that included only the three rounds of mentorships facilitated by MoH and laboratory implementing partners such as US CDC and MDH.

### Data analysis

Microsoft excel was used for data cleaning and analysis. We presented the descriptive analysis with frequency distributions (%) for categorical variables, the t-test was used to test the significance differenced for generic SLMTA program and modified SLMTA approach. The p-value of <0.05 was defined as significance differences between the two SLMTA program performance.

## Ethical considerations

This manuscript does not include human data, it did not require a formal review by Ethical Review Committees. The map that shows the distribution of accredited laboratories were drawn by QGIS desktop version 3.24.2 software. The approval to publish was granted by Ministry of Health through letter reference no. MA.155/174/01/625.

## Results

### SLIPTA assessment score of the SLMTA enrolled laboratories

Out of 138 SLMTA laboratories (S1 file) assessed by 2023 using the SLIPTA checklist, 81 (58.7%) scored 3 stars and above were enrolled in the accreditation process (Table 2). Of the 81 enrolled laboratories, 50 (61.7%) achieved ISO 15189 accreditation status by the year 2023. There was a significant increase in number of accredited laboratories during the five years (2018-2023) of modified SLMTA compared to the eight years (2010-2018) of generic SLMTA (47/64 vs. 3/17, *p*= 0.021). In the context of level of healthcare delivery, most of the accredited laboratories (n=47/50, 94%), belonged to the district and above (Table 3).

**Table 2.** SLIPTA assessment score of the SLMTA enrolled laboratories (2010-2023)

| Star level | Stars cut-off percentage | Number of laboratory | Proportion |
| --- | --- | --- | --- |
| 0 Star | <55 | 6 | 4.4 |
| 1 Star | 55-64 | 18 | 13.0 |
| 2 Stars | 65-74 | 33 | 23.9 |
| 3 Stars | 75-84 | 70 | 50.7 |
| 4 Stars | 85-94 | 9 | 6.5 |
| 5 Stars | $\geq 95$ | 2 | 1.5 |
| **Grand Total** |  | **138** | **100** |

**Table 3.**
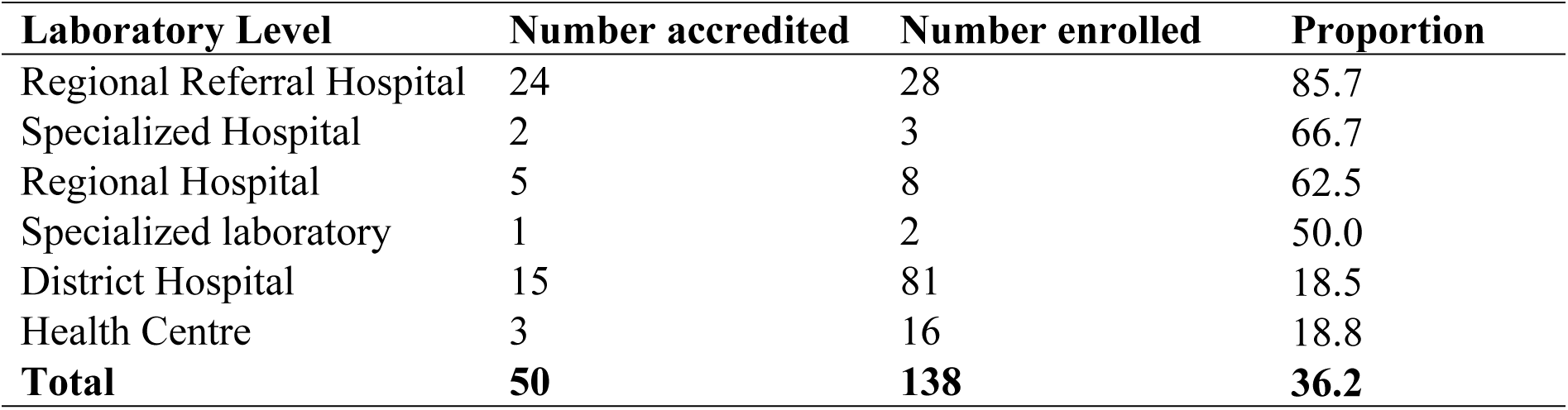
Proportion of accredited laboratories by level of service delivery (2010-2023)

### Trend of SLMTA accredited laboratories from 2010 to 2023

Since its adoption, and through the eight years of the SLMTA generic program implementation (2010-2018), only three out of the 17 (17.6%) enrolled laboratories into SLMTA program attained the status of three stars and above and eventually got accredited. In contrast, 47 out of 64 (73.4%) laboratories were accredited during the five years (2018-2023) of implementing a modified SLMTA approach (2018-2023); demonstrating a significant increase of 55.8% of accredited laboratories (*p*=0.021) (Fig 3).

**Fig 3.**
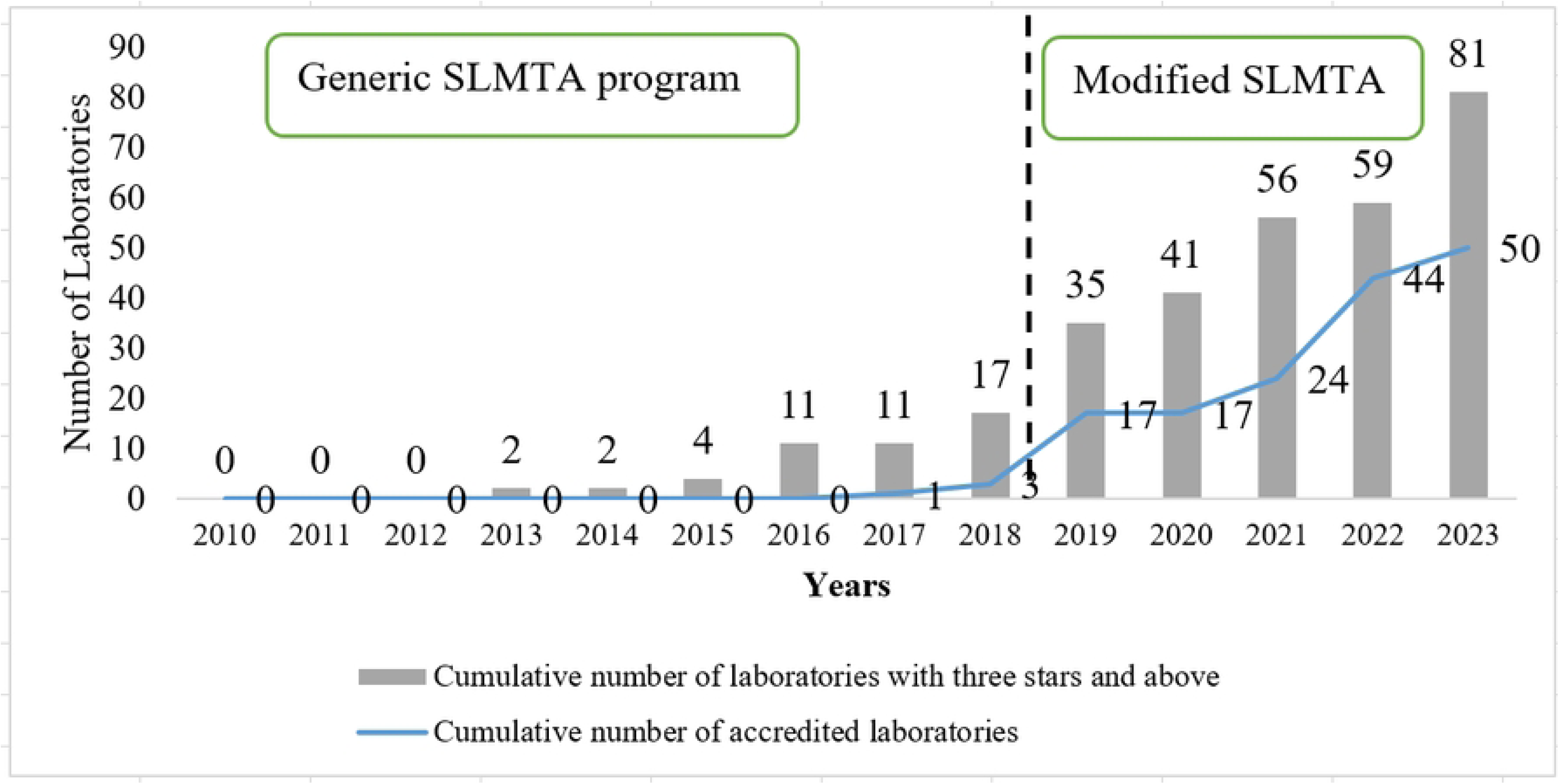
Accredited laboratories as per SLMTA enrollment criteria of three stars and above (2010-2023)

### Distribution of SLMTA accredited laboratories

The SMLTA accredited laboratories covered 23 (88.5%) of the 26 regions of Tanzania Mainland. Each of the 23 regions had at least one accredited laboratory with an exception of the Dar es Salaam region that had the highest number of 9 (18.0%) accredited laboratories (Fig 4).

**Fig 4.**
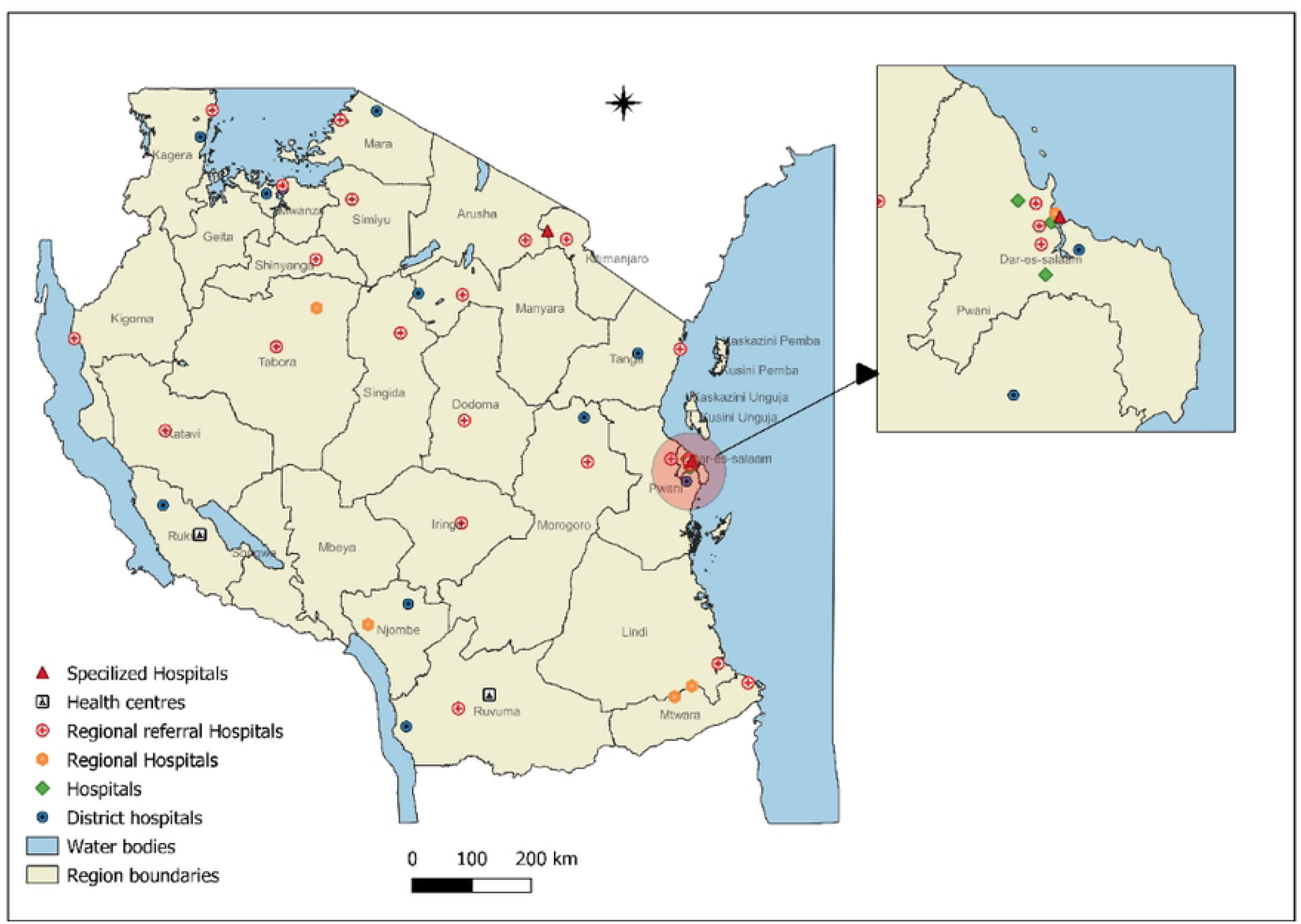
Distribution of SLMTA accredited laboratories by regions (2010-2023) in Tanzania Mainland. The map shows 26 Tanzania mainland administrative regions. Map were created using QGIS desktop 3.24.1. All shape files are openly available sources (https://www.nbs.go.tz/index.php/en/census-surveys/gis/385-2012phc-shapefiles-level-one-and-two). The shapefiles were made based on the 2012 population and housing census, but in this study, the shapefile has been modified to capture all the regions and district information.

### Accredited scope of the respective departments of the SLMTA laboratories

The three most common accredited scope were form Serology 43 laboratories, Tuberculosis 38 laboratories and Parasitology 25 laboratories. The Serology department had accredited tests that included Rapid tests for HIV, Syphilis (anti-Treponema antibodies), and Hepatitis B surface antigen (HBsAg), and Hepatitis C Virus. The Tuberculosis department had the tests scope covering MTB/Rif GeneXpert and AFB smear microscopy test. The three least accredited departments were: Immunology, Bacteriology, and Hematology (Fig 5).

**Fig 5.**
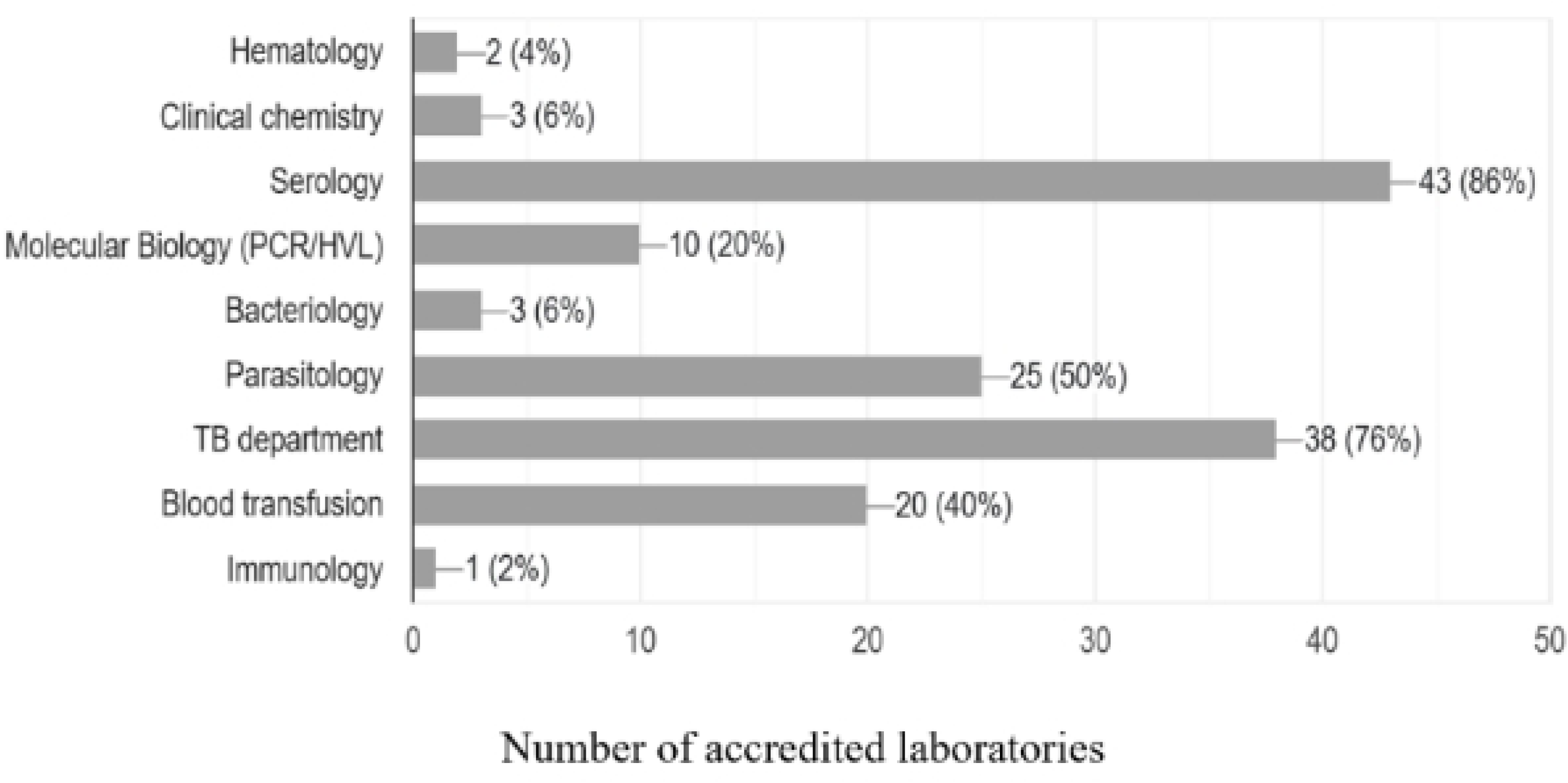
Accredited scope per laboratory departments.

## Discussion

Having accredited laboratories is the best way to regain the loss of trust in diagnostic services as the accreditation status attract international recognition of results and dictates quality (14). Guided therapy is critical in ensuring the proper management of patients to avoid unnecessary exposure to unwanted drug effects and irrational expenditure of resources (15). We are describing the Tanzania’s 13 years of remarkable revolutions in laboratory QMS through the implementation of the SLMTA program. The modifications made in the generic SLMTA approach resulted into a rise in number of accredited laboratories, from 3 in 2018 to 50 in 2023.

Implementation of the SLMTA-SLIPTA program using trained mentors and certified assessors certified has shown to be an effective way of attaining best results as documented in other African countries such as Ethiopia and Kenya (16,17). Like in the later countries, Tanzania managed to build the in-country pool of competent mentors and certified assessors with technical assistance from CLSI and ASLM. As reported earlier in other parts of Africa, training and mentorship programs have been a game-changer in attaining plausible results in augmentation of efforts towards strengthening of laboratory QMS to achieve ISO 15189 accreditation (18). Notably, in our 13-year’s journey, we noted significant achievements when the training and mentorship programs were tailored to address the key gaps that were identified during the evaluation of the generic SLMTA approach, eight years post its existence. Mastering the skills and competence of performing root cause analysis, corrective action, internal audit and method verification right at the start of the modified SLMTA made mentees more confident while working on their improvement projects independently.

Moreover, modification and customization of the training and mentorship models to suit the local context in our country was a catalyst for enhancing the accreditation process. This experience has also been shown to positively impact the outcome laboratory accreditation process in other African settings (19). The customized template documents.

Consistency and reproducibility of the procedures and results are one of the key factors in implementing QMS (20). This requires to have standard documents and records such as quality manuals, standard operating procedures, and laboratory register and request forms (21). As a strategy to save time and energy for document development, harmonization of QMS documents and development of the supportive supervision forms were done. This strategy not only brought uniformity to mentors and respective SLMTA implementing laboratories but also reduced the SLMTA program duration from a maximum of 18 months to nine months. Moreover, on the exit audits, laboratories that attained 3 stars or above were given the opportunity of being attached to an accredited laboratory (role model) to learn an effective approach towards accreditation within a short period. Similar strategy has been documented in Kenya, where the supported SLMTA laboratories were paired with internationally accredited research laboratories (19). On the hand, the development of the national Annual SLMTA and Accreditation Plan towards the end of every year of the modified SLMTA approach, served as the best monitoring, evaluation and learning tool for the action points.

Among many factors that contributed to the notable achievement of having more than 80% of SLMTA laboratories attaining 3 stars and above was the commitment of hospital leadership to support SLMTA implementation activities. Dedicated and committed mentors, qualified, skilled and motivated staff and the steady commitment of implementing partners across various regional areas have been very instrumental to the success of the attained accreditation status and efficiency of the SLMTA program as it was also reported in Kenya and other African countries (17).

The serology department, including rapid tests, had more accredited scope due to most of the serological tests being in vertical program which had consistent supply of reagents and consumables. Implementing partners focus on additional tests like HIV rapid tests and syphilis to achieve the UNAIDS 90-90-90 goal (22). Other frequently accredited scope included AFB smear microscopy and GeneXpert MTB/RIF tests for the TB program, whereas in the Malaria Program, tests like Malaria rapid diagnostic test (mRDT) and Malaria blood slides were among the most accredited scope at the parasitology department. The support and funding of all tests under the vertical program led to an increase in the scopes of laboratory accreditation (23). The observed improvements can be attributed to the enhanced availability of supplies and the functionality of laboratory equipment, which significantly influence the effectiveness, reliability, and timeliness of laboratory data (ISO 15189:2022). These factors collectively contribute to the overall improvement of laboratory services, ensuring accurate and dependable test results essential for healthcare delivery (24).

The selection of laboratories through an application process may have introduced bias, favouring those who felt ready, with more resources or better initial conditions. This might reflect the concentration of accredited laboratories in Dar es Salaam which is a major commercial city in Tanzania.

## Conclusion

To the best of our knowledge, this is the first report of the modified SLMTA approach with remarkable results. The modified SLMTA program significantly improved the status and number of accredited laboratories in Tanzania. While the attained milestone demonstrates the SLMTA program’s effectiveness in strengthening quality of laboratory services, the significant achievements underscore the essence of monitoring, evaluation and learning of the implemented programs, and most importantly the need for strategies that contextualize and tailor the program for sustainability.

## Supporting information

**S1 file**. SLMTA dataset 2023

## Data Availability

Dataset is available

## Acknowledgments

We are very grateful to the Ministry of Health, President’s Office, Regional Administration and Local Government (PO-LARG), Tanzania Field Epidemiology and Laboratory Training Program (FELTP) for their technical support on this study.

## Author Contributions

**Conceptualization:** Peter Richard Torokaa, Jacob Lusekelo, Charles Massambu and Nyambura Moremi.

**Data curation**: Peter Richard Torokaa, Jacob Lusekelo and Regnald Julius.

**Formal analysis:** Peter Richard Torokaa.

**Methodology:** Peter Richard Torokaa, Nyambura Moremi, Regnald Julius, Charles Massambu, Maria E. Kelly, Mtebe Majigo, Agricola Joachim, Alice Kyalo and Jacob Lusekelo.

**Resources:** Peter Richard Torokaa, Regnald Julius, Nyambura Moremi, Charles Massambu, and Jacob Lusekelo.

**Supervision:** Nyambura Moremi and Jacob Lusekelo.

**Validation:** Peter Richard Torokaa Regnald Julius, Alice Kyalo, and Jacob Lusekelo.

**Writing-original draft:** Peter Richard Torokaa, Nyambura Moremi, Regnald Julius, Charles Massambu, Maria E. Kelly, Mtebe Majigo, Agricola Joachim, Alice Kyalo and Jacob Lusekelo.

**Writing – review & editing:** Peter Richard Torokaa, Nyambura Moremi, Alex Sifael Magesa, Regnald Julius, Charles Massambu, Maria E. Kelly, Mtebe Majigo, Agricola Joachim, Alice Kyalo and Jacob Lusekelo.

